# Decreased DNA methylation at promoters and gene-specific neuronal hypermethylation in the prefrontal cortex of patients with bipolar disorder

**DOI:** 10.1101/2020.12.10.20246405

**Authors:** Miki Bundo, Junko Ueda, Yutaka Nakachi, Kiyoto Kasai, Tadafumi Kato, Kazuya Iwamoto

## Abstract

Bipolar disorder (BD) is a severe mental disorder characterized by repeated mood swings. Although genetic factors are collectively associated with the etiology of BD, the underlying molecular mechanisms, particularly how environmental factors affect the brain, remain largely unknown. We performed promoter-wide DNA methylation analysis of neuronal and nonneuronal nuclei in the prefrontal cortex of patients with BD (N=34) and controls (N=35). We found decreased DNA methylation at promoters in both cell types in the BD patients compared to the controls. Gene Ontology (GO) analysis of differentially methylated region (DMR)-associated genes revealed enrichment of molecular motor-related genes in neurons, chemokines in both cell types, and ion channel- and transporter-related genes in nonneurons. Detailed analysis further revealed that growth cone- and dendrite-related genes, including *NTRK2* and *GRIN1*, were hypermethylated in neurons of BD patients. To assess the effect of medication, neuroblastoma cells were cultured under therapeutic concentrations of three different mood stabilizers (lithium, valproate, and carbamazepine). We observed that up to 37.9% of DMRs detected in BD overlapped with mood stabilizer-induced DMRs. Interestingly, mood stabilizer-induced DMRs showed the opposite direction of changes in DMRs in BD, suggesting the therapeutic effects of mood stabilizers on DNA methylation. Among the DMRs, 12 overlapped with loci identified by a previous genome-wide association study of BD. Finally, we performed qPCR analysis of 10 DNA methylation-related genes and found that *DNMT3B* was overexpressed in BD. The cell type-specific DMRs identified in this study will be useful for understanding the pathophysiology of BD.

## Introduction

Bipolar disorder (BD), also known as manic depressive illness, is a severe and common mental disorder characterized by repeated mood swings of depressive and manic episodes, with elevated rates of mortaility^1, 2^. Early epidemiological and linkage studies suggested that BD is a highly heritable disorder caused by a complex interaction of genetic and environmental risk factors^3^. Genome-wide association studies (GWAS) revealed that BD is a polygenic disorder caused by multiple genetic risks with small effect sizes, similar to schizophrenia (SZ), and shared genetic risks with other psychiatric disorders, such as SZ and autism^4-6^. Additionally, the contribution of *de novo* loss-of-function mutations, as well as *de novo* copy number variations, has been suggested in the etiology of BD^7-9^. Although the genetic landscape of BD is gradually becoming understood, heritability estimated from epidemiological studies is modestly accounted for by these genetic studies.

Epigenetics, including DNA methylation, reflects gene-environment interactions during development and affects long-lasting gene expression status in cells^10^. Therefore, unraveling the epigenetic landscape of psychiatric disorders will contribute to the understanding of the heritability and pathophysiology of psychiatric disorders^11-13^. In BD, several candidate-gene-based approaches have been performed, such as *BDNF, COMT*, and *SLC6A4* genes in postmortem brains^12^. Additionally, comprehensive DNA methylation studies have revealed expression-linked DNA methylation changes in the cerebellum^14^, accelerated aging in the hippocampus^15^, loss of brain laterality associated with *TGFB2* methylation^16^, and methylation imbalance of synaptic function-related genes between the frontal and temporal cortices^17^. However, there have been no established findings that were replicated in multiple studies.

DNA methylation status in brain cells shows great variation among cell types^18-20^. Therefore, cell type-specific epigenetic analysis will also be important. Recent studies have highlighted that cell type-specific epigenetic differences are linked to SZ and neuropsychiatric traits^21-24^. In BD, hypomethylation of the *IGF2* enhancer, which is associated with increased tyrosine hydroxylase protein levels, has been reported in isolated neuronal nuclei^25^.

In this study, we performed promoter-wide DNA methylation analysis of neuronal and nonneuronal nuclei in the prefrontal cortex (PFC) of patients with BD. In addition to identifying cell type-specific differentially methylated regions (DMRs), we found hypomethylation at promoters in both cell types in BD patients. The affected genes included hypomethylation of molecular motor-related genes in neurons, chemokine-related genes in both cell types, and ion channel- and transporter-related genes in nonneurons. We also found neuron-specific hypermethylation of growth cone- and dendrite-related genes. We then assessed the effect of medication by using neuroblastoma cells and found that up to 37.9% of DMRs in BD patients overlapped with mood stabilizer-induced DMRs. Interestingly, mood stabilizer-induced DMRs showed the opposite direction of changes in DMRs in BD, suggesting the therapeutic effects of mood stabilizers on DNA methylation. Among the DMRs, 12 overlapped with loci identified by a previous GWAS of BD^6^. We also found overexpression of *DNMT3B* in BD and SZ, suggesting possible molecular mechanisms of neuronal hypermethylation.

## Materials and Methods

### Postmortem brains

PFC (Brodmann area 46) samples of patients with BD (N = 34) and controls (N = 35) were obtained from the Stanley Medical Research Institute. Among them, selected samples were also analyzed by reduced representation bisulfite sequencing (RRBS). Demographic variables are summarized in **Table S1**. This study was approved by the ethics committees of participating institutes (the Research Ethics Committee of Kumamoto University, the Research Ethics Committee of the Faculty of Medicine of The University of Tokyo, the Ethical Review Board of Juntendo University, and the Wako 1st Research Ethics Committee of RIKEN).

### Nuclei preparation

Neuronal and nonneuronal nuclei fractions were separated by NeuN-based cell sorting^18^. In brief, after homogenization of fresh-frozen brain samples, the nuclear fraction was retrieved by Percoll discontinuous density gradient centrifugation. An anti-NeuN antibody (#MAB377, Millipore, Burlington, MA, USA) conjugated with Alexa Fluor 488 was used for staining. NeuN+ and NeuN− nuclei were sorted using the FACS Aria system (BD Biosciences, Franklin Lakes, NJ, USA) as previously described^18^.

### Cell culture

Cell culture and drug conditions were as previously described^26^. In brief, cells of the human neuroblastoma cell line SK-N-SH (American Type Culture Collection) were cultured for 8 days in Eagle’s minimal essential medium containing 10% fetal bovine serum with one of three mood stabilizers. The medium was changed on days 2, 5, and 8. On day 9, the cells were retrieved. We used three mood stabilizers: lithium chloride (Sigma-Aldrich; St. Louis, MO, USA), valproic acid sodium salt (Sigma-Aldrich), and carbamazepine (Sigma-Aldrich). Valproate and lithium were directly dissolved into the medium. Carbamazepine was dissolved in dimethyl sulfoxide before it was added to the medium. The concentration of each drug was determined based on the therapeutic concentrations for BD patients. We prepared the following minimum and maximum therapeutic concentrations: valproate, 0.3 mM and 0.6 mM; lithium, 0.6 mM and 1.2 mM; and carbamazepine, 0.05 mM and 0.1 mM.

### Enrichment of methylated DNA and tiling arrays

Enrichment of methylated DNA was performed using MethylCollector (Active Motif, Carlsbad, CA, USA) according to the manufacturer’s protocol. A total of 100 ng of DNA was used, and methylated DNA was retrieved in 100 µL of elution buffer. In qPCR, aliquots of eluted methylated DNA were used for quantification. Probe preparation and labeling for Affymetrix human promoter 1.0R tiling arrays were performed according to the Affymetrix chromatin immunoprecipitation assay protocol (Affymetrix, Santa Clara, CA, USA) as previously described in detail^18^. The array covers 25,500 human promoters by 4.6 million 25-mer oligo probes. Each promoter covers approximately 7.5 kb upstream through 2.5 kb downstream of the transcription start site by 35 bp probe spacing.

### Data analysis

In the postmortem brain experiment, all experiments were performed in duplicate using independently prepared probes (experiments 1 and 2). The total numbers of array data points were therefore 136 and 140 for BD patients and controls, respectively, which have been deposited in the Gene Expression Omnibus as GSE137921. References were prepared by applying human genomic DNA amplified by a GenomiPhi V2 DNA amplification kit (GE Healthcare, Chicago, IL) to MethylCollector. The number of methylated regions (MRs) of each sample was counted by MAT software^27^ using two replicate sample datasets (experiments 1 and 2) as one target group and a reference dataset (whole-genome amplified samples) as a reference group. The number of MRs was compared using the Mann-Whitney test. Principal component analysis (PCA) of MRs based on Jaccard statistics was conducted using bedtools^28^. DMRs were independently identified in experiments 1 and 2 by comparing the patient and control datasets. The DMRs detected in experiments 1 and 2 were then intersected by bedtools and used for further analysis.

The parameters used in the MAT were as follows: bandwidth, 300 bp; max gap, 300; min probe, 10; P-value, 1e-3. The MRs and DMRs on the sex chromosomes were excluded from this analysis. DMRs within multigene families, such as olfactory receptors and protocadherins, were removed from the analysis. Annotation was conducted using HOMER^29^. DMR-associated genes were additionally identified if the distance between the intergenic DMR and the nearest TSS was less than 5 kb. Gene Ontology (GO) analysis was performed with ToppGene^30^. The chromosomal location of the DMRs was visualized using CHARANGO software^31^.

### RRBS

RRBS was performed using the MethylSeq library construction, sequencing, and data analysis service (Zymo Research, Irvine, CA, USA). The total number of RRBS data points was 10 each for the BD and controls, which has been deposited in the DDBJ Sequence Read Archive as DRA008934. In brief, a total of 500 ng of genomic DNA was digested with Taq I and Msp I. DNA fragments were filled in, and A was added at the 3’-end, followed by adaptor ligation. After bisulfite modification using the EZ DNA methylation-Direct kit (Zymo Research), preparative-scale PCR was performed. Size selection was performed on a 4% NuSieve 3:1 agarose gel. Library material was recovered using a Zymoclean Gel DNA recovery kit (Zymo Research). DNA sequencing was performed using an Illumina HiSeq2000. The data were analyzed using a Zymo Research proprietary pipeline (Zymo Research). We used CpG sites 1) with coverage ≥ 10 and 2) located within DMRs detected in the array analysis, which corresponded to 4,959 and 3,281 CpG sites for neuronal and nonneuronal DMRs, respectively. Statistical analyses were conducted by a one-tailed Student’s t-test.

### Quantitative RT-PCR

Real-time quantitative RT**-**PCR was performed according to a previous study^32^. A total of 1 µg of total RNA was used for cDNA synthesis by oligo (dT) and SuperScript II reverse transcriptase (Invitrogen). qRT**-**PCR was performed using SYBR/GREEN I dye (Applied Biosystems, Foster City, CA, USA) with ABI PRISM 7900HT (Applied Biosystems). The comparative Ct method (Applied Biosystems) was employed for quantification. We used two internal control genes based on our previous analysis^32^. In addition to analyzing all the measured samples, we performed pH-adjusted analysis because the brain sample pH systematically affects the transcriptome^33^. Low pH samples (pH < 6.4) were omitted from the pH-adjusted analysis. The threshold was previously determined^34^. P < 0.05 in the Mann-Whitney test was considered significant. Primer pairs used in this study were as follows: MBD1, 5’-AGGAGGACAAGGAGGAGAACAA-3’ and 5’-GGCTGAAAATCTCCGTGATCAC-3’; MBD2, 5’-TCCAGGCAGAACCAATCCTTTC-3’ and 5’-AAAAGACATGGTCCCTGCCCT-3’; MBD2L, 5’-GTTTGGCTTAACACATCTCAACCC-3’ and 5’-GTACTCGCTCTTCCTGTTTCCTGA-3’; MBD3, 5’-GCTCCCTGTCAGAGTCAAAGCAC-3’ and 5’-GCACCAACCTCAGGAAGACGT-3’; MBD4, 5’-AATGGACACCTCCTCGGTCACC-3’ and 5’-CTTCCAAAGCACAGGTATTGCC-3’; DNMT1, 5’-CACTGCACGTGTTTGCTCC-3’ and 5’-ACCCGAGCTCAACCTGG-3’; DNMT2, 5’-TATGCGGTGACATGGATGAAC-3’ and 5’-TCTCATCACCCCAATCAGAAAC-3’; DNMT3A, 5’-AACCTTCCCGGTATGAACAGGC-3’ and 5’-TGCTGAACTTGGCTATCCTGCC-3’; DNMT3B, 5’-CCGTGACTGCAATAGAACCCTC-3’ and 5’-AGAACTCAGCACACCCCTTCCT-3’; MECP2, 5’-GCCTCCTTTCCGTTTGATTTG-3’ and 5’-CACATTGAGTAACAGTCCTGGTGA-3’. For each primer pair, amplification of the single product was confirmed by gel electrophoresis and by monitoring the dissociation curve.

### Data availability

The array data are available under accession GSE137921. The RRBS data are available under accession DRA008934.

## Results

### Widespread promoter hypomethylation of the PFC in patients with BD

We performed promoter-wide DNA methylation analysis on NeuN-sorted neuronal (NeuN+) and nonneuronal (NeuN-) nuclear fractions derived from the PFC of patients with BD (N = 34) and controls (N = 35) (**Table S1**). DNA fragments containing densely methylated CpGs were enriched using the MBD2B/3L complex and analyzed with a promoter tiling array. PCA of the DNA methylation signature revealed a clear separation between neurons and nonneurons (**Fig. 1a**). We then compared the total number of MRs per sample (**Fig. 1b**). Consistent with our previous report^18^, the total number of MRs was significantly lower in neurons than in nonneurons within controls (P = 0.0006) and within patients (P = 4.75E-05). Additionally, a significant decrease in the total number of MRs was identified in both neurons and nonneurons of patients compared to controls (P = 0.0031 and P = 0.0318, respectively, in the Mann-Whitney test). The decreased number of MRs in the patients was not dependent on the genomic context, such as repeat structure or segmental duplications (**Fig. S1**), implying genomic context-independent, promoter-wide hypomethylation in the cells of the PFC of patients with BD.

**Figure 1.**
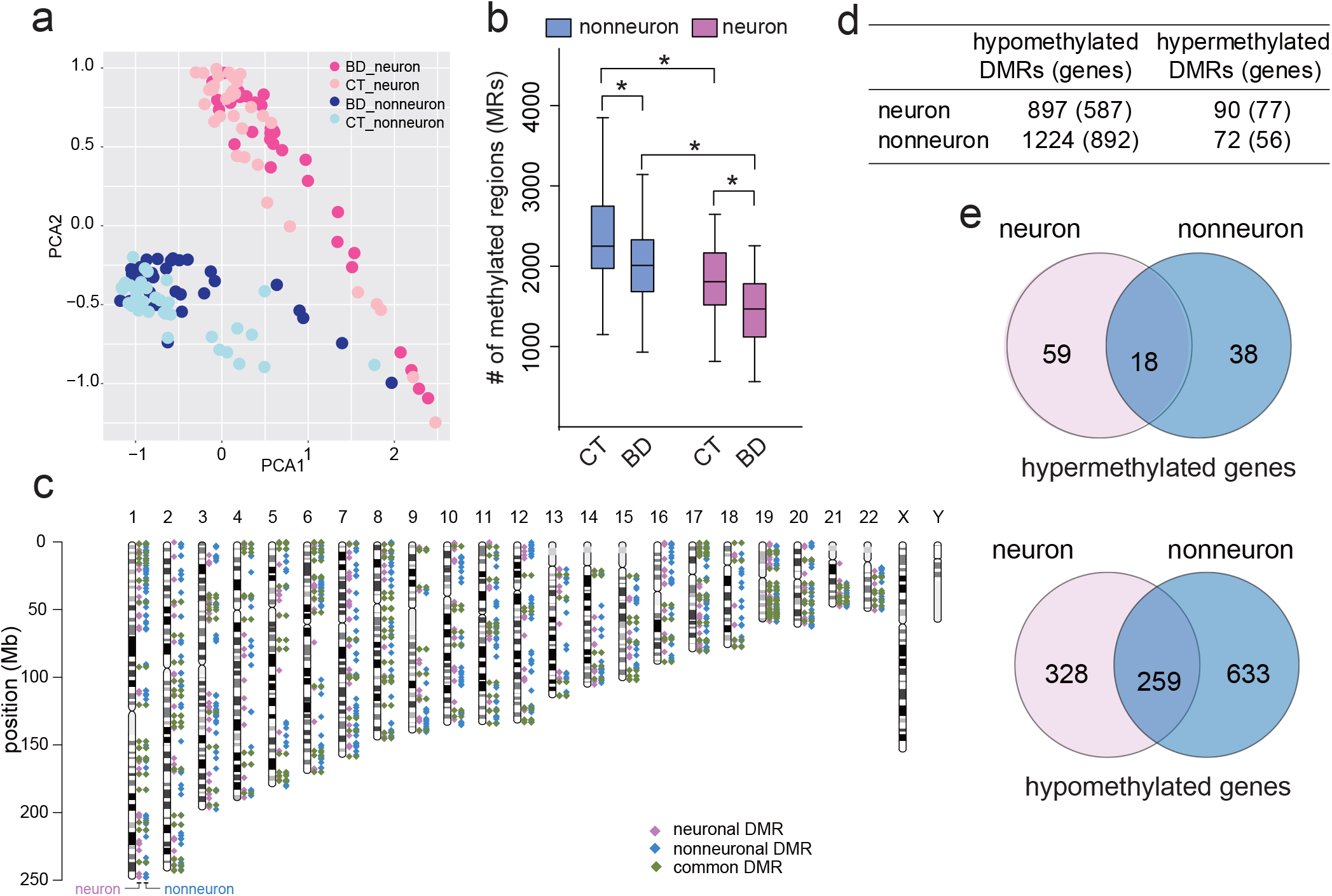
MRs and DMRs in BD. a. PCA of the MRs of each sample. b. Decreased number of MRs in BD. * shows significant changes by the Mann-Whitney test. c. Chromosomal locations of DMRs. d. Total number of DMRs and DMR-associated genes. e. Venn diagrams of DMR-associated genes. Detailed information on DMRs is shown in **Tables S2** and **S3**. MR, methylated region; DMRs, differentially methylated regions; BD, bipolar disorder; PCA, principal component analysis; CT, control.

### Identification and characterization of DMRs

We then identified DMRs between BD patients and controls in each cell type (**Tables S2 and S3**). The DMRs of neurons and nonneurons are uniformly distributed throughout the genome (**Fig. 1c**). Consistent with the decreased number of MRs in BD, most DMRs in both neurons and nonneurons showed hypomethylation (**Fig. 1d**). The overlaps between neurons and nonneurons at the gene level ranged from 23.4% to 44.1%. The rest showed cell-type-specific DNA methylation changes (**Fig. 1e**). We performed GO analysis using all the DMR-associated genes (**Fig 2a**). Each of the significantly enriched GO terms was composed of neuronal and nonneuronal DMR-associated genes at different ratios (**Fig. 2a**). We found that kinesin complex-, microtubule-, and motor molecule-related genes dominantly included neuronal DMR-associated genes, whereas chemokine activity-related and inflammation-related genes were evenly enriched among both neuronal and nonneuronal DMR-associated genes. In contrast, ion channels and transporter-related terms mainly included nonneuronal DMR-associated genes. To further extract the cell-type-specific signature, we performed stratified GO analysis considering cell type and direction of methylation change (**Fig. 2b, Table S5**). Strikingly, hypermethylated genes in neurons included genes related to the growth cone and dendrites (**Fig. 2b**), such as the NMDA NR1 subunit gene *GRIN1* and the BDNF receptor gene *NTRK2* (**Fig. 2c**). Both genes have been the long-studied genes in psychiatric disorders, and their downregulation in the postmortem brains of BD patients has been established (see Discussion). On the other hand, kinesin complex- and microtubule motor activity-related genes were included in the hypomethylated genes of neurons (**Fig. 2b, Table S5**).

**Figure 2.**
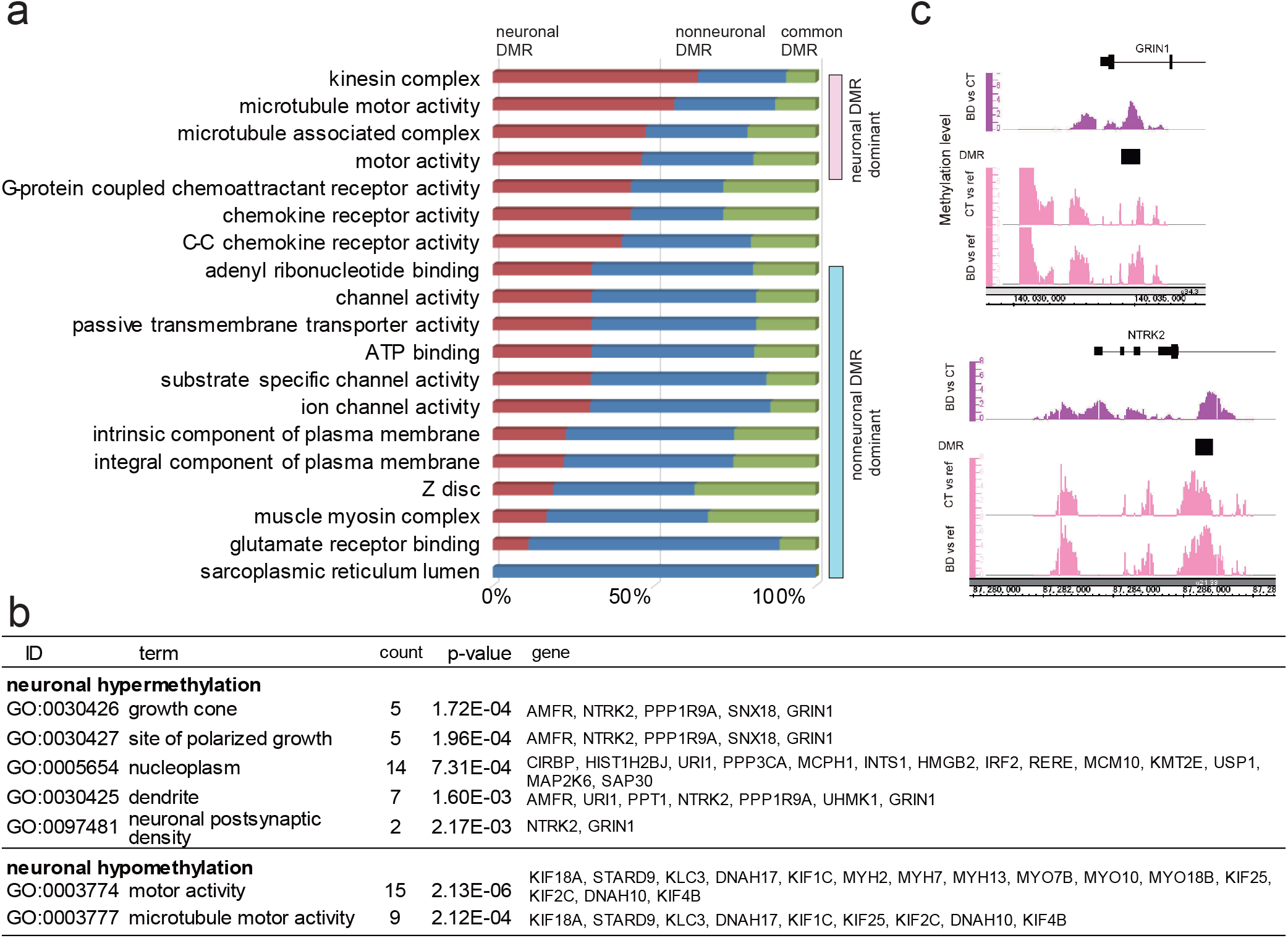
GO analysis of DMRs. a. GO analysis of all DMR-associated genes. Significant GO terms were sorted based on the ascending order of the percentage of NeuN+ DMR-associated genes. See **Table S4** for detailed results. b. GO analysis of DMR-associated genes considering the cell type and direction of methylation changes. Only the results of neuronal DMR-associated genes are presented. See **Table S5** for the results of nonneurons. c Example of neuronal hypermethylation at *GRIN1* and *NTRK2*. DMRs are denoted by black squares. The top panel shows DNA methylation levels determined by direct comparison of BD (N = 34) and CT (N = 35) samples. The peaks for the target group (BD) are shown to highlight the DMR. The bottom panels show average DNA methylation levels determined by comparison of either CT (N = 35) or BD (N = 34) with references (nonmethylated human genome). GO, gene ontology; DMRs, differentially methylated regions; CT, control; BD, bipolar disorder; MR, methylated region.

### Technical considerations of MRs and DMRs by RRBS and qPCR

We performed RRBS analysis in neurons and nonneurons of the selected subjects (**Tables S1** and **S6**). Approximately 95% of the MRs detected in the array showed greater than 70% of the DNA methylation levels in RRBS, ensuring high sensitivity to the detection of methylated DNA (**Fig. S2**). A total of 999 DMRs contained at least one CpG site whose DNA methylation level could be determined by RRBS. Among them, 190 DMRs contained CpG(s) showing significant DNA methylation differences by RRBS (**Fig. S2**). The average validation rates by RRBS were 16.4% for hypomethylation and 52.7% for hypermethylation. Hypermethylation changes were more supported by RRBS than hypomethylation changes. Because all the arbitrarily chosen hypomethylated DMRs were successfully confirmed by independent qPCR (**Fig. S3**), the low rate of replication of hypomethylated DMRs by RRBS may support the involvement of other epigenetic regulations, such as hmC^35^ (see Discussion). Based on the DMRs confirmed by RRBS, typical DNA methylation differences were estimated to range from 12.4 to 17.8% (**Table S7**).

### Assessment of the effect of mood stabilizers

We then assessed the effect of mood stabilizers on DNA methylation changes using a human neuroblastoma cell line. Cells cultured under the minimum and maximum therapeutic concentrations of three different mood stabilizers for 8 days were retrieved, and their DNA methylation patterns were profiled with the same array platform (**Fig. 3a**). We examined the relationship between the DMRs detected in BD patients and those detected in cell culture. We found that 31.3% and 37.9% of the neuronal and nonneuronal DMRs, respectively, overlapped with DMRs detected in at least one cell culture condition (**Fig. 3b, Tables S2** and **S3** for details of overlapping DMRs). Regarding the direction of methylation changes in the DMRs, both directions showed a similar extent of overlap (**Fig. 3b**). Further analysis revealed that hypomethylated DMRs in BD patients showed a greater overlap with hypermethylated DMRs in cell culture and *vice versa* among both neuronal DMRs (**Fig. 3c**) and nonneuronal DMRs (**Fig. 3d**).

**Figure 3.**
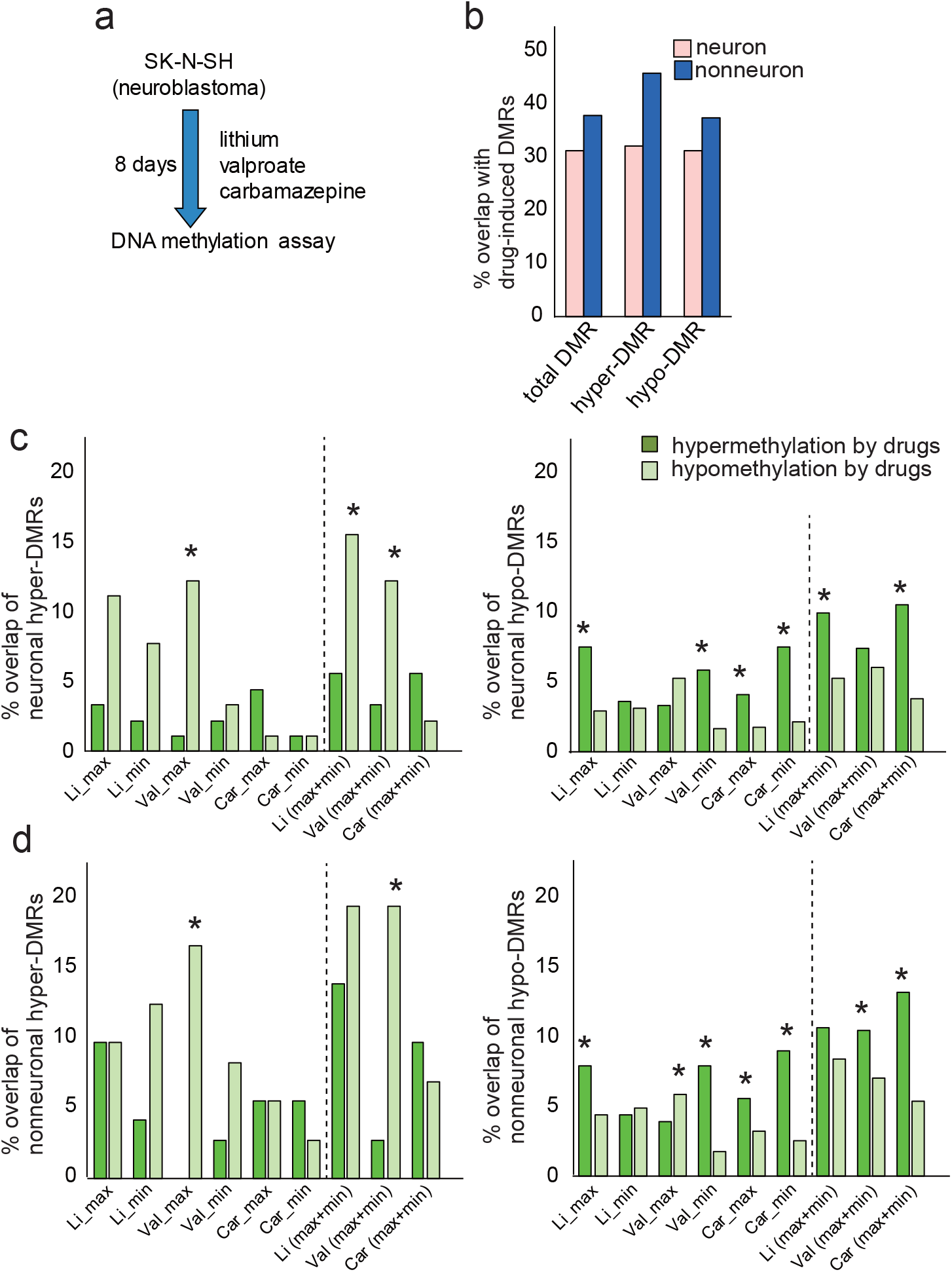
Effect of mood stabilizers on DMRs. a. Experimental scheme for cell culture. b. % overlap with drug-induced DMRs. Detailed analysis of the overlap between either neuronal DMRs (c) or nonneuronal DMRs (d) and drug-induced DMRs. The abbreviations max, min, and (max+min) indicate maximum, minimum, and maximum or minimum therapeutic concentrations of mood stabilizers. * indicates a significant difference in Fisher’s exact test (P < 0.05). Detailed information on overlapping DMRs is shown in **Tables S2** and **S3**. DMRs, differentially methylated regions; BD, bipolar disorder; Li, lithium; Val, valproate; Car, carbamazepine.

### Overlap analysis with a GWAS of BD

We compared the chromosomal loci identified by a GWAS of BD with the DMRs. Among the 30 loci identified by the largest GWAS of BD^6^, 8 loci included a total of 12 DMRs (**Fig. 4a**). At the gene level, we also identified additional overlapping genes between the results of the GWAS and those of this study, including *CACNA1C, SHANK2*, and *GRIN2A* (**Fig. 4b**).

**Figure 4.**
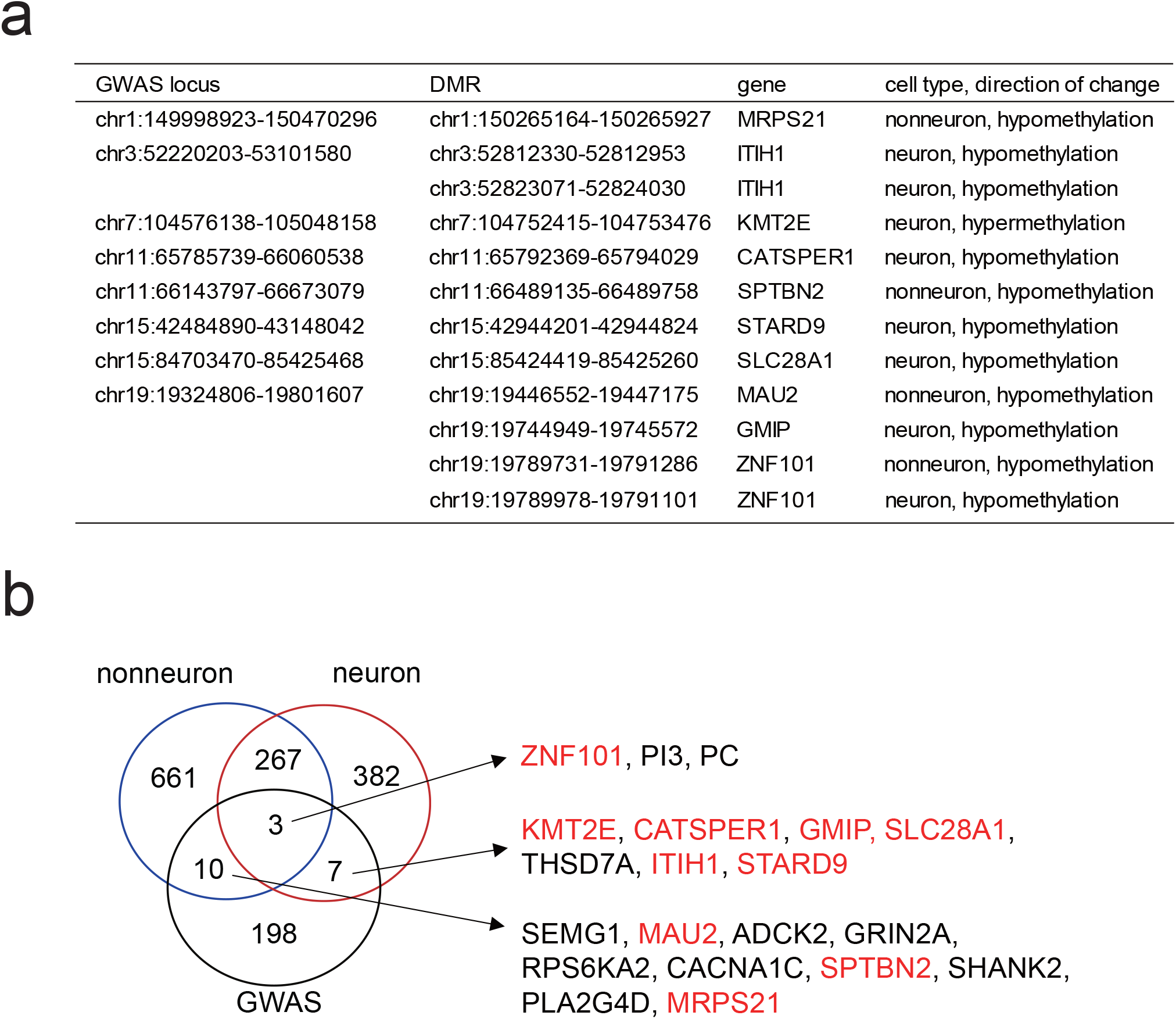
The overlap between GWAS loci and DMRs. a. Overlap at the chromosomal locus level (hg19). b. Overlap at the affected gene level. Genes indicated in red show that those DMRs are included in the GWAS loci. Note that the total number of DMR-associated genes shows some inconsistencies with Figure 1 due to the presence of genes having multiple DMRs of both directions of changes. GWAS loci and genes were retrieved from Stahl *et al*^6^. GWAS, genome-wide association study; DMRs, differentially methylated regions.

### qPCR of DNA methylation-related genes

To examine the genes involved in the DNA methylation changes in BD, we measured the gene expression levels of 4 DNA methyltransferases and 5 methyl-CpG binding domain-containing proteins by qRT-PCR using bulk PFC samples (**Fig. 5a**). Among the measured genes, the expression of *DNMT3B*, which is highly expressed in neurons, showed an increase compared to that in controls by using the multiple internal control genes for normalization and by pH-adjusted analyses (**Fig. 5b**). Specific and increased expression of *DNMT3B* was also found in the PFC of SZ patients (**Fig. 5b**), suggesting that *DNMT3B* is involved in altered DNA methylation in psychosis.

**Figure 5.**
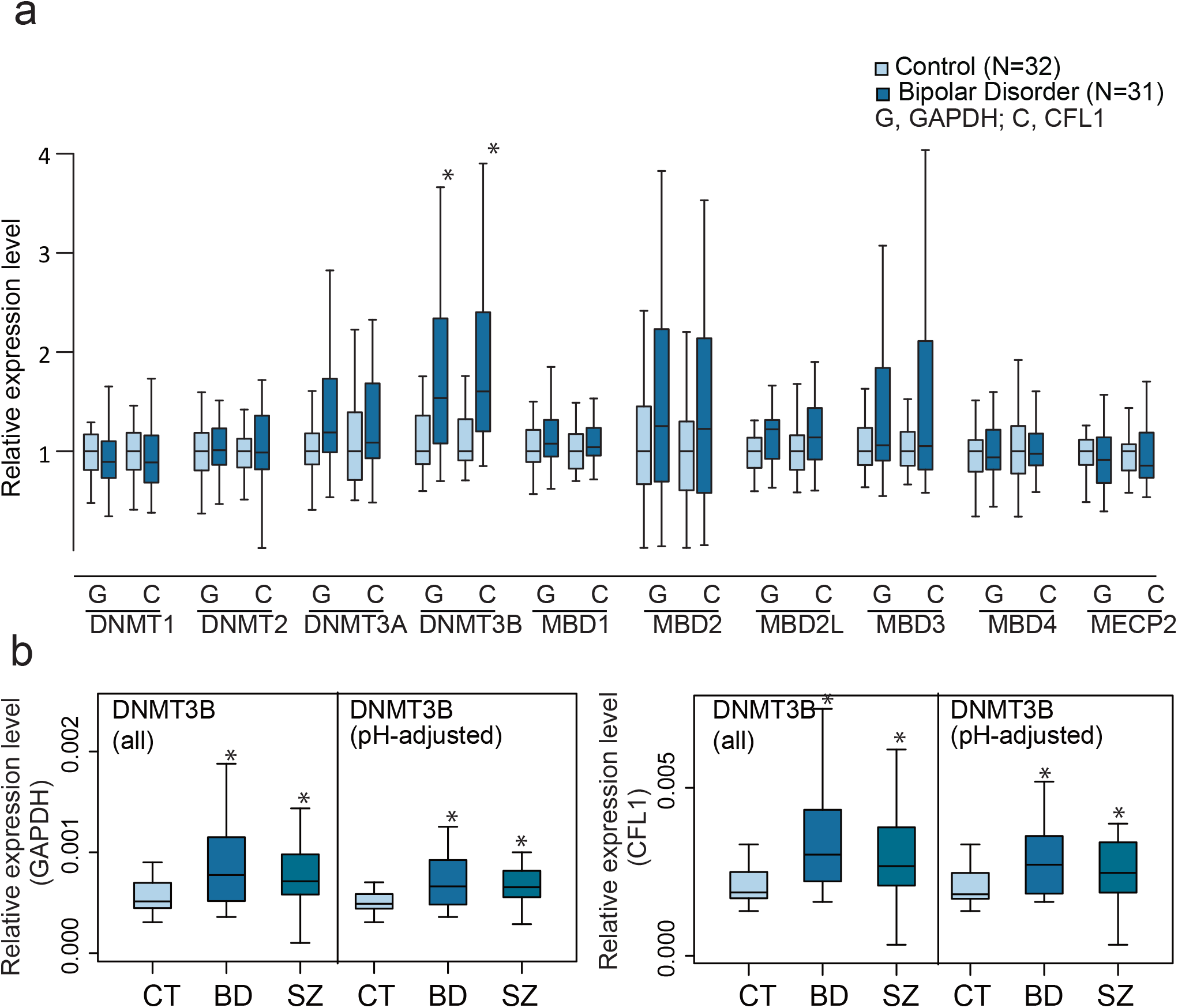
qRT-PCR analysis of DNA methylation-related genes. a. Expression levels of DNA methylation-related genes. Expression levels were plotted relative to the average value of the control. Note that for some subjects, total RNA samples were not available. b. The expression level of *DNMT3B* in BD and SZ. The expression levels of two different genes, GAPDH: G and CFL1: C, were used as internal controls. In the pH-adjusted analysis, samples for which the brain sample pH was below 6.4 were removed from the analysis based on a previous study.^34^ * indicates a significant change in the Mann-Whitney test compared to the CT (P < 0.05). CT, control; BD, bipolar disorder; SZ, schizophrenia

## Discussion

Here, we performed brain cell type-specific DNA methylation analysis on the PFC of BD patients. Our analysis revealed a tendency towards decreased promoter methylation of both neurons and nonneurons and neuronal hypermethylation in some key genes important for neuronal function in BD.

We employed the enrichment of methylated DNA by MBD2B/3L followed by promoter tiling array analysis. Compared to the bisulfite sequencing (BS)-based method, this approach has limitations in the coverage of the genome and accuracy of quantitative determination. However, taking advantage of the binding specificity of MBD2B, which does not bind hydroxymethylcytosine (hmC)^35^, we were able to enrich and analyze the MRs consisting of methylcytosine (mC)^36, 37^. Excluding hmC would be particularly important because hmC is enriched in neurons and cannot be discriminated from mC by the BS^19^, which was supported by the failure to replicate hypomethylated sites by bisulfite sequencing in this study. Therefore, the DMRs defined based on only the mC will be valuable for interpretation of epigenetic signatures in the brain. By performing promoter-wide analysis, we focused on the genomic regions directly important for gene expression regulation. Genome-wide analysis, such as MBD-Seq, would be useful for future studies to understand the entire role of epigenetic regulation in BD. Another limitation would be that due to the enrichment-based method, it cannot estimate the methylation level by calculating the ratio of methylated to unmethylated signals. However, the accuracy of quantification has also been proven in MBD-Seq by using the appropriate reference set^36^. Although we could not adopt such correction in this study, we determined DMRs by independent duplicate assays, and we estimated that the DMRs showed substantial DNA methylation changes by RRBS analysis of the selected samples (**Table S7**).

### DNA hypomethylation in BD

In control brains, we replicated the decreased number of neuronal MRs compared to nonneuronal MRs^18^. On the other hand, previous BS analyses by others reported higher methylated levels in neurons than in nonneurons^19, 23^. Discordance may come from differences in the data interpretation involving higher hmC levels in neurons and in the genomic region analyzed in this study, i.e., promoters in this study and the entire genomic region in other studies. hmC may also affect the lower validation rate of hypomethylated DMRs by RRBS (**Table S7**). The global tendency toward hypomethylation and gene-specific neuronal hypermethylation in BD was seemingly contradictory. Although the molecular mechanism and relationship between these changes are unclear, such changes have also been observed in cancer cells^38^.

Global DNA hypomethylation is frequently reported in blood cells of psychiatric disorders, including BD^39-41^. We also reported global DNA hypomethylation in BD and SZ in blood cells, and hypomethylation was associated with the serum level of the methyl-group donor betaine (*N,N,N*-trimethylglycine)^42^. Whether hypomethylation in the PFC of BD patients is accompanied by similar metabolite changes remains unclear. Systematic DNA methylation changes across different tissues have been reported during normal aging, enabling the establishment of the epigenetic clock^43^. Interestingly, BD patients showed accelerated aging in blood and brain tissues^44,45^. It would be interesting to pursue the relationship between promoter-wide hypomethylation and accelerated aging in BD.

### DMR-associated genes

By GO analysis, we found that motor activity-related terms were enriched in DMR-associated genes. Most of them showed hypomethylated changes in neurons (**Fig. 2b**). Genes included seven kinesin complex genes (*KIF*s and *KLC3*), six myosin components (*MYH*s and *MYO*s), one lipid transfer protein (*STARD9*), and two dynein complexes (*DNAH17* and *DNAH10*), suggesting that motor molecules in neurons are widely dysregulated. Because neurons must transport synaptic vesicle precursors, neurotransmitter receptors, and mRNAs over long distances^46^, dysregulation of motor activity affects diverse neuronal functions and the pathophysiology of psychiatric disorders. Interestingly, altered microtubule functions in neural stem and mature neural cells in BD have been recently reported^47^.

We found hypermethylation of growth cone- and dendrite-related genes. Among them, *NTRK2* and *GRIN1* have been the long-studied genes in psychiatric disorders, and their downregulation in the postmortem brains of BD patients has been established. *NTRK2*, also known as TrkB, encodes a BDNF receptor and has been one of the prime targets in mood disorders. Decreased expression of *NTRK2* was repeatedly reported in postmortem brains of patients with psychiatric disorders^48-51^ and animal models of depression^52-54^. The BDNF-NTRK2 signaling pathway is critical for the antidepressant effect of lamotorigine^55^ and ketamine^56, 57^ as well as the antimanic effect of lithium^58^ in animal models. Genetic studies have revealed that *NTRK2* is associated with the treatment response to mood stabilizers in BD^59, 60^ and suicidal behavior in mood disorders^61^. Interestingly, hypermethylation of the CpG island of the *NTRK2* promoter has been reported in suicide completers^62^. Because the identified region in this study was close to the CpG island, these methylation changes may be linked and contribute to the pathophysiology of psychiatric disorders.

NMDA receptors (NMDARs) mediate basic neuronal functions, and their dysfunction is closely linked to the pathophysiology of psychiatric disorders^63^. *GRIN1* (*NR1*) encodes an essential subunit of NMDAR, and its downregulation was reported in the postmortem brains of patients with psychiatric disorders^64^. *GRIN1* knockdown mice showed a wide range of behavioral alterations related to psychiatric disorders^65^. The involvement of altered DNA methylation of NMDAR genes, including *NR1*, which is associated with changes in expression and subunit composition, has been reported^66-68^

Other characteristic findings include a wide range of DNA methylation changes in ion channel- and transporter-related genes in nonneurons. For example, they include four calcium channels (*CACNA1C, CACNA2D4, CACNB2*, and *CACNG8*), three glutamate receptors *(GRIK2, GRIN2A*, and *GRM8*), four GABA receptors (*GABRA5, GABRG3, GABRG3*, and *GABRP*), and nine potassium channels (*KCNA1, KCNA4, KCNA7, KCNAB2, KCNAB2, KCND3, KCND3, KCNE1*, and *KCNG2*). A recent GWAS showed enrichment of GWAS signals in calcium signaling genes and genes expressed in neurons^69^. The current finding of DNA methylation changes in ion channels, including calcium channels, sheds light on the potential roles of these channels in nonneurons, such as oligodendrocytes, microglia, and astrocytes, in BD. Further analysis of the specific nonneuronal cell population will be important.

### Effect of mood stabilizers on epigenetic alterations

We observed that up to 37.9% of DMRs in BD overlapped with mood stabilizer-induced DMRs in cultured cells (**Fig. 3b**). Despite the simple cell culture model, these overlapping DMRs and opposite directions of changes between the postmortem brain and cell culture imply the pathophysiological importance of these DMRs. A similar opposite direction of DNA methylation changes related to mood stabilizers has been reported not only in a gene-specific manner^26^ but also in systematic alterations in accelerated aging in BD^70^. Although the precise molecular mechanism remains unclear, mood stabilizers can normalize epigenetic regulation in brain cells^12^, leading to the amelioration of multiple DMRs between BD patients and controls.

### Comparison with GWAS results

At the chromosomal location level, among the DMRs overlapping between GWAS and this study, we regarded *KMT2E* and *SPTBN2* as particularly important (**Fig. 4a**). *KMT2E* encodes histone lysine methyltransferase 2E. Loss of function of histone lysine methyltransferases is involved in BD, SZ, and autism^8, 71, 72^, and cell-type-specific alteration of histone lysine modification in postmortem brains and animal models of psychiatric disorders has been reported^73^. *SPTBN2*, also known as *SCA5*, regulates glutamate signaling by stabilizing EAAT4, and mutations in *SPTBN2* cause spinocerebellar ataxia type 5^74^. At the gene level, several genes overlapped with GWAS, including the well-studied genes in BD^6^ such as *CACNA1C, SHANK2*, and *GRIN2A* (**Fig. 4b**). They seemed to appear as candidates due to their long gene length. Taken together, epigenetic alterations of GWAS-implicated genes provide important insights into the molecular pathophysiology of BD. The relationship between risk SNPs and DNA methylation status will be an important issue to be studied.

### Overexpression of *DNMT3B* in BD and SZ

The role of DNA methylation-related genes, especially *DNMT1*, in BD and SZ has been well studied^12^. In this study, we found an increase in *DNMT3B* expression in both BD and SZ, implicating the possible role of *DNMT3B* in neuronal hypermethylation in BD (**Fig. 5**). Whether patients with SZ show epigenetic changes similar to those of patients with BD needs to be studied. Increased expression of *DNMT3B*, but not *DNMT1* and *DNMT3A*, was recently reported in learned helplessness rats, supporting its role as a stress-inducible, psychiatric disorder-related neuronal DNA methyltransferase^75^. However, the molecular mechanism underlying *DNMT3B* overexpression and gene-specific neuronal hypermethylation remains unknown. Additionally, the examination of genes responsible for the cytosine modification pathway, such as the TET family, base-excision repair genes and methyl-donor metabolism-related genes, would be considered candidate genes potentially involved in epigenetic dysregulation in BD.

## Conclusion

We observed cell-type-specific, pathophysiology-related DNA methylation changes in the PFC of patients with BD and identified increased expression of *DNMT3B* as a potential molecular mechanism. The present findings may help in understanding the neurobiological mechanisms underlying bipolar disorder.

## Acknowledgments

This work was supported in part by the UTokyo Center for Integrative Science of Human Behavior (CiSHuB) and by the International Research Center for Neurointelligence (WPI-IRCN) at The University of Tokyo Institutes for Advanced Study (UTIAS). Postmortem brains were donated by the Stanley Microarray Collection, courtesy of Drs Michael B. Knable, E. Fuller Torrey, Maree J. Webster, and Robert H. Yolken. We are indebted to the Research Resource Center at the RIKEN for nuclear sorting and microarray analysis. We would like to thank Taeko Miyauchi and Fumiko Sunaga for their technical assistance.

