## supplementary figures for "Decreased DNA methylation at promoters and gene-specific neuronal hypermethylation in the prefrontal cortex of patients with bipolar disorder"

**Suppplementary Figures**

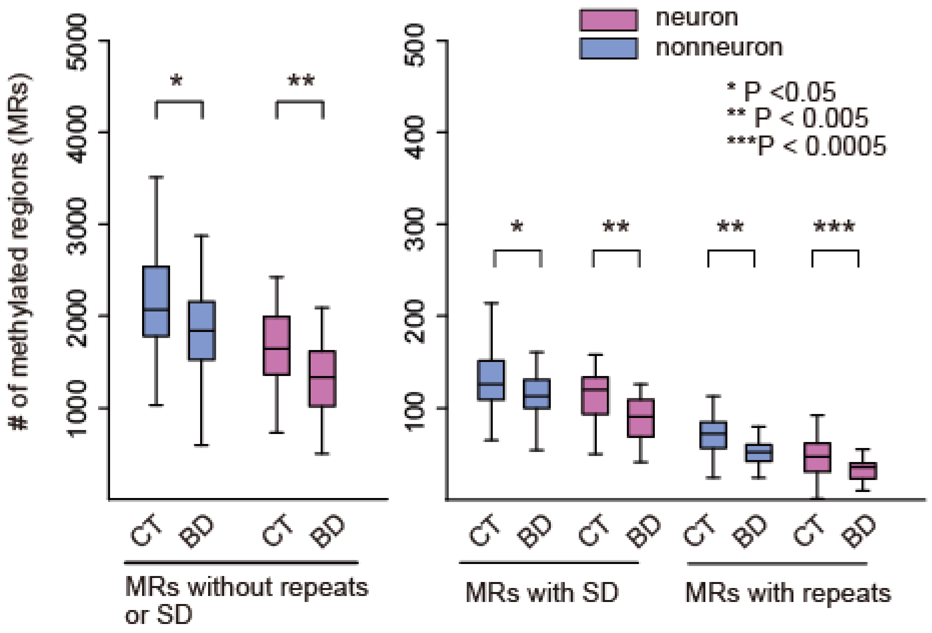


**Figure S1.** Decreased number of MRs in BD. Left panel: comparison of the average number of MRs excluding the MRs that overlapped with repetitive sequences or segmental duplications. Right panel: comparison of the average number of MRs that overlapped with repetitive sequences or segmental duplication. Note that the decreased number of MRs in BD was not influenced by the cell type or genomic context. CT, control; BD, bipolar disorder; MR, methylated region; SD, segmental duplication.

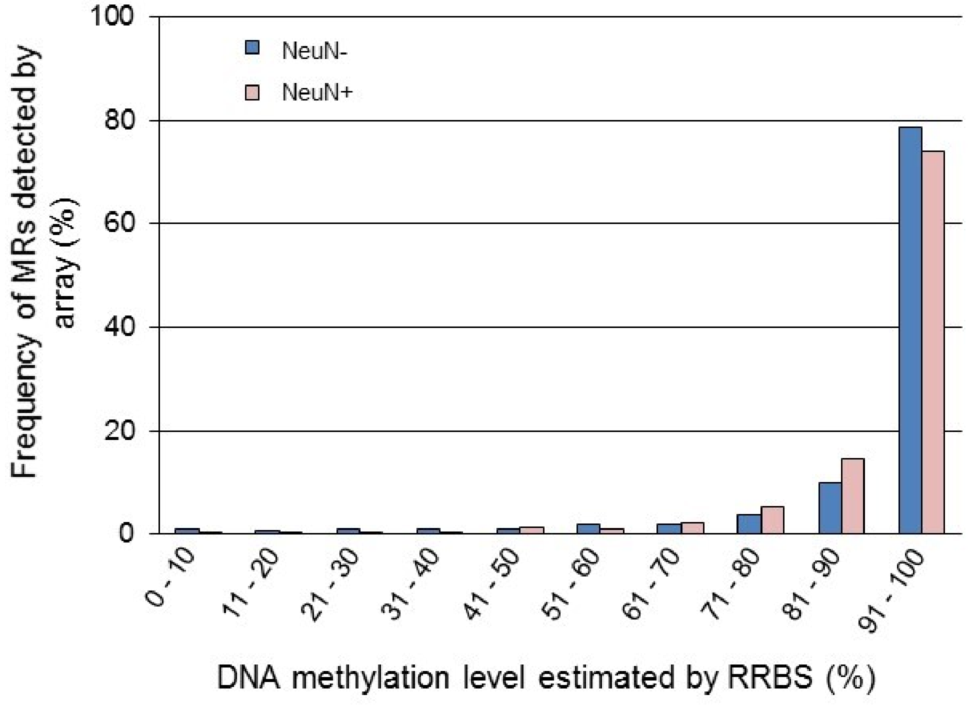


**Figure S2. MRs identified by array analysis were efficiently detected by RRBS**. For each MR detected by array, the average DNA methylation level estimated by RRBS was calculated. Approximately 95% of MRs were estimated to have greater than 70% of the DNA methylation level in both NeuN+ and NeuN- samples. DNA methylation levels were calculated using the CpGs with coverage greater than 10. A representative result (from one subject) is shown here.

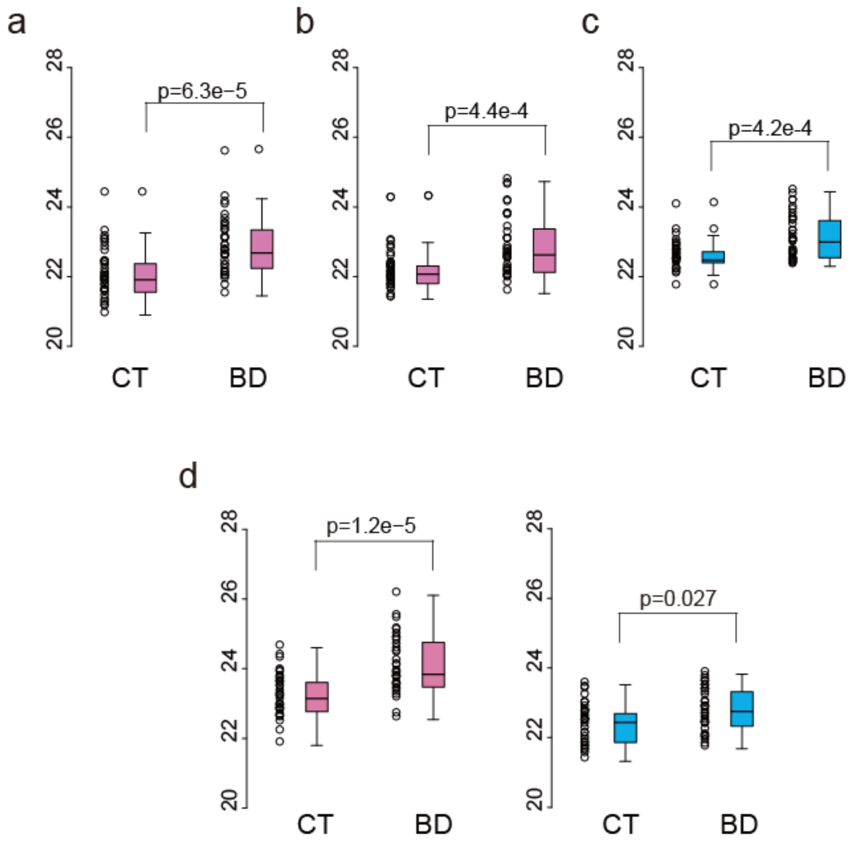


**Figure S3. qPCR validation of hypomethylated regions in BD.** a. Neuronal hypomethylation at chr8:6478908-6480044 in *MCPH1*. b. Neuronal hypomethylation at chr4:57459500-57460123 in *THEGL*. c. Nonneuronal hypomethylation at chr19:49932808-49933573 in *SLC17A7*. d. Neuronal and nonneuronal hypomethylation at chr2:136593910-136594677 in *LCT*. For qPCR, aliquots of eluted methylated DNA were used for quantification. The CT value of each sample was plotted. The P values were obtained by the Mann-Whitney test.
